# From Omics Data to Biological Context: The CURIE AI Engine. Illustrated with the Transcriptomic Response to Bacterial Pneumonia

**DOI:** 10.64898/2026.10.19.26365885

**Authors:** Giulia Castellani, Alejandro Rodriguez-Mena, Xavier Tarragó-Claramunt, Victor Madarnás-Gómez, Yovani Torres-Favier, Javier Méndez-Viera, Antonio Monleón-Getino

## Abstract

High-throughput sequencing necessitates robust computational tools to contextualize complex transcriptomic data. In this study, we analyzed RNA-Seq data from peripheral blood of bacterial pneumonia patients and healthy controls (GEO: GSE161731) to investigate systemic molecular responses to infection. Differential gene expression analysis identified significantly upregulated genes in pneumonia. We used GANGO for Gene Ontology (GO) term enrichment and BioFunctional for hierarchical exploration, providing an initial overview of affected biological processes. To deepen interpretation, we leveraged CURIE (www.curieai.tech), an AI platform that cross-references GO terms with scientific literature. CURIE generates narrative explanations, linking molecular mechanisms, experimental evidence, and publications to each biological process, moving beyond simple enrichment. Our combined analysis revealed a coordinated upregulation of pathways involved in DNA repair and replication, transcriptional activation, RNA processing, translation, ribosome biogenesis, mitochondrial respiration, and cofactor binding. By integrating these findings within a broad, literature-supported framework, we highlighted how multiple cellular compartments and processes collaborate to mount an effective response to infection-induced stress. This study underscores the potential of combining classical statistical enrichment with AI-based interpretation for a holistic understanding of host-pathogen interactions at the transcriptomic level.

## 1. Introduction

The advent of high-throughput RNA sequencing (RNA-Seq) has transformed how we explore transcriptomic landscapes, making it possible to study gene expression changes with a level of detail that was unthinkable until recently. This has opened up new ways to investigate how complex biological systems respond to different challenges, including infections. Understanding the molecular shifts that happen during bacterial infections is key to uncovering the mechanisms behind host defense, tissue adaptation, and disease progression. These insights can ultimately support the development of targeted therapies and improve clinical management strategies.

Traditionally, RNA-Seq studies rely on differential expression analysis followed by functional enrichment to highlight biological processes and pathways that stand out among the regulated genes. While these methods are powerful for making sense of large datasets, they often fall short of providing the broader context needed to truly understand the biological meaning of the changes observed.

To bridge this gap, more recent approaches have started to combine statistical enrichment with advanced computational tools that help place the findings into a wider biological perspective. In this study, we analyzed transcriptomic data from peripheral blood samples of individuals with bacterial pneumonia to get a clearer picture of the systemic molecular responses triggered by infection. After identifying differentially expressed genes and their enriched functions, we used CURIE (Rodriguez-Mena et al., 2025; Tarragó et al, 2025a; Tardiolo et al., 2026), an AI platform built to mine scientific literature and automatically generate explanatory summaries. By merging these AI-driven insights with standard analyses, our goal was to build a more complete and biologically meaningful interpretation of how the host transcriptional program responds to bacterial pneumonia.

## 2. Materials and Methods

### 2.1. RNA-Seq Data Acquisition and Preprocessing

The RNA-Seq dataset analyzed in this study was retrieved from the public repository GEO under accession number GSE161731. This dataset originates from the study titled “Dysregulated transcriptional responses to SARS-CoV-2 in the periphery” and includes transcriptomic profiles from peripheral blood samples of individuals diagnosed with bacterial pneumonia, SARS-CoV-2, seasonal coronavirus, influenza, and healthy controls.

Raw count data and associated sample metadata were downloaded and filtered to retain only human genes annotated as protein-coding, using the biomaRt package and the Ensembl GRCh38 reference (dataset: hsapiens_gene_ensembl). Samples corresponding to the Bacterial and Healthy cohorts were selected for comparative analysis.

Differential gene expression analysis was performed using the DESeq2 package (Love et al., 2014). The DESeq2 workflow included normalization of read counts and statistical testing with the DESeq() function. Genes with an adjusted p-value (padj) < 0.1 were considered statistically significant. These genes were further stratified by their log2 fold change (log2FC): Genes with log2FC > 0 were classified as upregulated in the Bacterial group while genes with log2FC < 0 were classified as upregulated in the Healthy group.

The final output was a structured table containing ensembl gene ID, differential group assignment (Bacterial or Healthy), and organism taxonomy (Homo sapiens). This table served as the input file for the enrichment analysis.

### 2.2. Functional Analysis Pipeline: GANGO and BioFunctional

Prior to this research work focused on the use and validation of CURIE, a bioinformatics pipeline was developed, described, and presented with a complete use case in “GANGO + BioFunctional: A Computational Tool for Efficient Functional Gene Analysis” (Rodrigez-Mena et al., 2025). GANGO + BioFuncional, an R-based Shiny application designed for end-users, addresses these challenges by providing a streamlined and comprehensive workflow for functional gene analysis. This interactive and freely available tool requires no installation, thus significantly enhancing its accessibility. The application is composed of two primary modules: GANGO, which efficiently processes input data and performs functional annotation to Gene Ontology (GO) terms and KEGG pathways; and BioFuncional, which is dedicated to in-depth analysis and interpretation.

The list of differentially expressed genes was analyzed using GANGO, a computational algorithm developed to facilitate ontology and pathway mapping in multi-group datasets. GANGO processes gene or taxonomic identifiers and assigns each to corresponding Gene Ontology (GO) terms and KEGG pathways through functional annotation.

The core operations of GANGO included mapping Ensembl IDs to GO and KEGG terms using curated annotation databases, retaining group-level metadata to enable differential analysis across cohorts, and enrichment analysis to assess over-representation of ontologies in each group, generating EA (Enrichment Analysis) values.

The output of GANGO is a comprehensive table linking each gene to relevant GO terms and group assignments. This served as the input for BioFunctional.

BioFunctional is an interactive Shiny-based R application designed for the hierarchical exploration and visualization of gene function enrichment results. It processes the output of GANGO and provides tools for advanced filtering, classification, and graphical representation of biological ontologies.

The table generated by GANGO was filtered and subjected to functional analysis within the BioFunctional module. The process began with data preprocessing, during which the GANGO output was reformatted and cleaned, and the essential columns for downstream analysis and biological interpretation were selected. Following this, a hierarchical annotation of Gene Ontology (GO) terms was performed. Using the QuickGO API, ancestral ontology information was retrieved for each GO term, allowing the integration of hierarchical relationships. Enrichment significance was assessed by transforming the Enrichment Analysis (EA) values into standardized Z-scores. This normalization allowed for the comparison of GO terms on a unified scale.

Finally, the BarPlot function of BioFunctional was used to visualize the Gene Ontology (GO) terms associated _93_ with bacterial pneumonia that displayed the highest enrichment Z-scores. These were categorized by the GO do- _94_ main, specifically highlighting Biological Process (Figure 1), Molecular Function (Figure 2), and Cellular Compo- _95_ nent (Figure 3).

**Figure 1:**
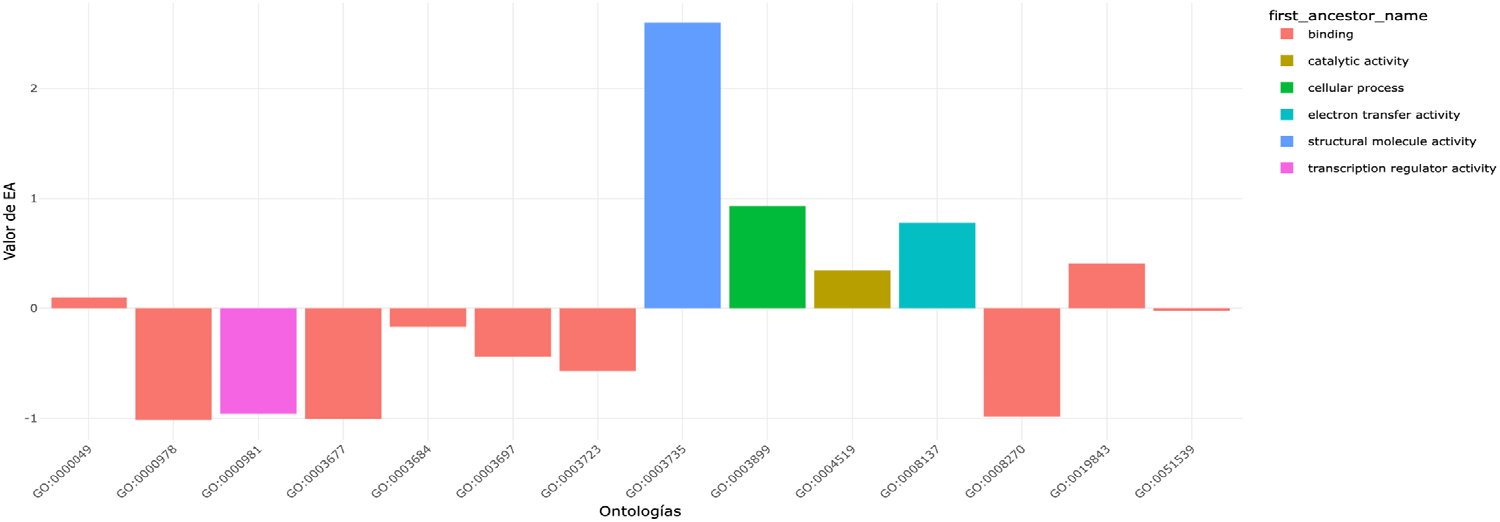
GO belonging to the Molecular function group.

**Figure 2:**
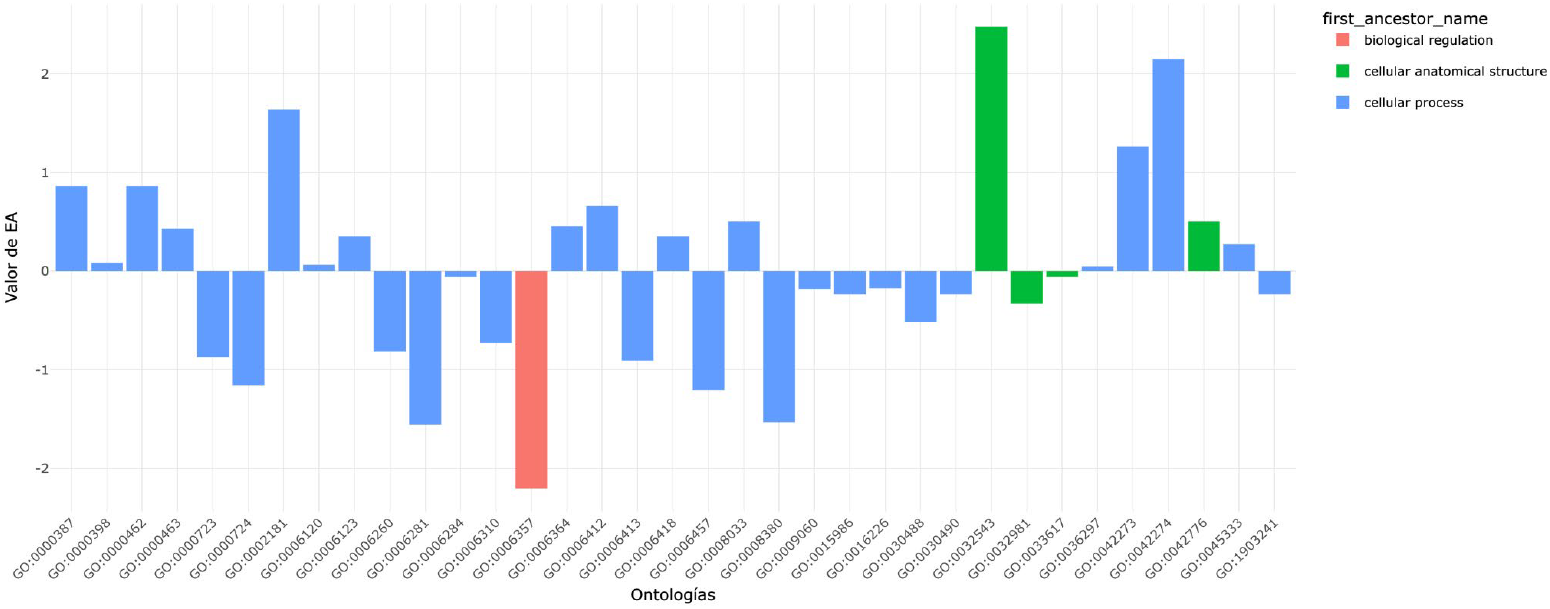
GO belonging to the Biological process group.

**Figure 3:**
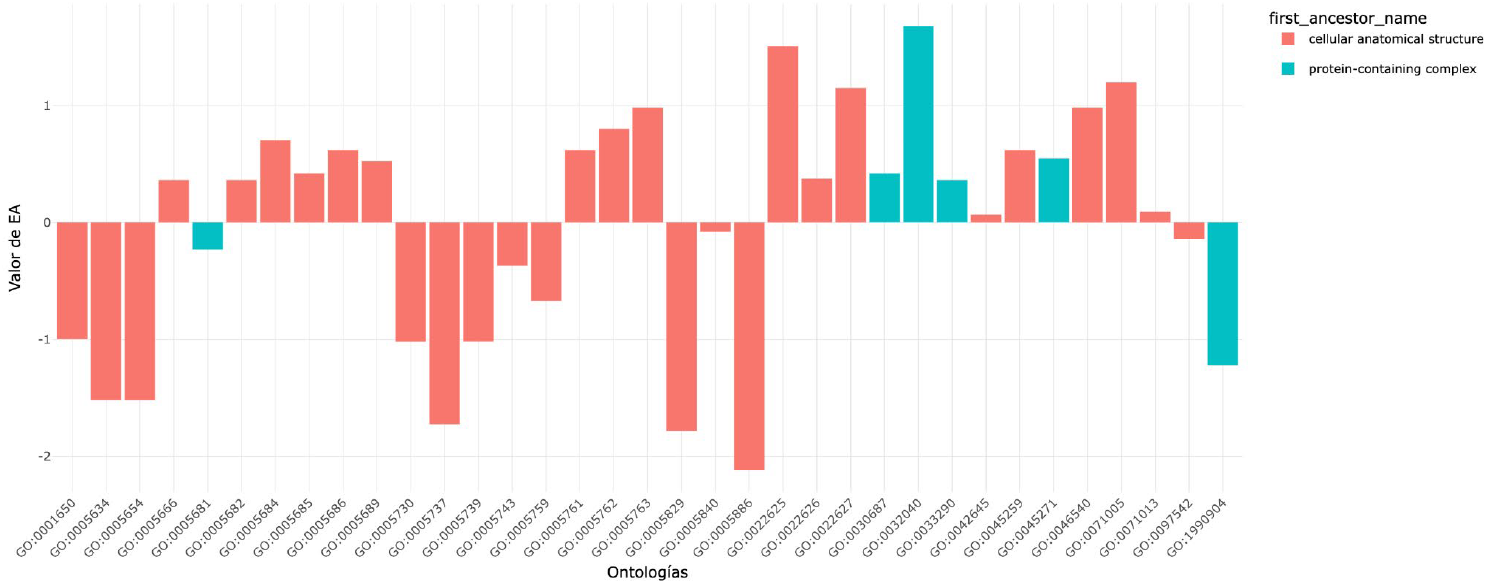
GO belonging to the Cellular component group.

The most significantly enriched GO terms identified through this visualization were subsequently compiled into a new table (Table 1). In this table, the terms were further grouped based on the biological processes in which they are involved. Only the GO up-regulated in the Bacterial group, as compared to the healthy controls, were included.

**Table 1:** Significantly Enriched GO Terms Upregulated in the Bacterial Group Compared to Healthy Controls. This table presents Gene Ontology (GO) terms that were significantly enriched and subsequently grouped by their involvement in biological processes. Only GO terms upregulated in the bacterial group, relative to the healthy controls, are included.

| GO | Name | Section |
| --- | --- | --- |
| GO:1903241 | U2-type prespliceosome assembly | Transcription and RNA Processing |
| GO:0071005 | U2-type precatalytic spliceosome | Transcription and RNA Processing |
| GO:0071013 | catalytic step 2 spliceosome | Transcription and RNA Processing |
| GO:0046540 | U4/U6 x U5 tri-snRNP complex | Transcription and RNA Processing |
| GO:0030488 | tRNA methylation | Transcription and RNA Processing |
| GO:0008380 | RNA splicing | Transcription and RNA Processing |
| GO:0008033 | tRNA processing | Transcription and RNA Processing |
| GO:0006364 | rRNA processing | Transcription and RNA Processing |
| GO:0006357 | regulation of transcription by RNA polymerase II | Transcription and RNA Processing |
| GO:0005689 | U12-type spliceosomal complex | Transcription and RNA Processing |
| GO:0005685 | U1 snRNP | Transcription and RNA Processing |
| GO:0005686 | U2 snRNP | Transcription and RNA Processing |
| GO:0005682 | U5 snRNP | Transcription and RNA Processing |
| GO:0005684 | U2-type spliceosomal complex | Transcription and RNA Processing |
| GO:0005681 | spliceosomal complex | Transcription and RNA Processing |
| GO:0005666 | RNA polymerase III complex | Transcription and RNA Processing |
| GO:0003899 | DNA-directed RNA polymerase activity | Transcription and RNA Processing |
| GO:0003723 | RNA binding | Transcription and RNA Processing |
| GO:0000978 | RNA polymerase II cis-regulatory region sequence-specific DNA binding | Transcription and RNA Processing |
| GO:0000387 | spliceosomal snRNP assembly | Transcription and RNA Processing |
| GO:0000398 | mRNA splicing, via spliceosome | Transcription and RNA Processing |
| GO:0000049 | tRNA binding | Transcription and RNA Processing |
| GO:0042274 | ribosomal small subunit biogenesis | Translation and Ribosome Biogenesis |
| GO:0042273 | ribosomal large subunit biogenesis | Translation and Ribosome Biogenesis |
| GO:0033290 | eukaryotic 48S preinitiation complex | Translation and Ribosome Biogenesis |
| GO:0032543 | mitochondrial translation | Translation and Ribosome Biogenesis |
| GO:0030490 | maturation of SSU-rRNA | Translation and Ribosome Biogenesis |
| GO:0032040 | small-subunit processome | Translation and Ribosome Biogenesis |
| GO:0030687 | preribosome, large subunit precursor | Translation and Ribosome Biogenesis |
| GO:0022626 | cytosolic ribosome | Translation and Ribosome Biogenesis |
| GO:0022625 | cytosolic large ribosomal subunit | Translation and Ribosome Biogenesis |
| GO:0022627 | cytosolic small ribosomal subunit | Translation and Ribosome Biogenesis |
| GO:0019843 | rRNA binding | Translation and Ribosome Biogenesis |
| GO:0006418 | tRNA aminoacylation for protein translation | Translation and Ribosome Biogenesis |
| GO:0006413 | translational initiation | Translation and Ribosome Biogenesis |
| GO:0006412 | translation | Translation and Ribosome Biogenesis |
| GO:0005840 | ribosome | Translation and Ribosome Biogenesis |
| GO:0005763 | mitochondrial small ribosomal subunit | Translation and Ribosome Biogenesis |
| GO:0005761 | mitochondrial ribosome | Translation and Ribosome Biogenesis |
| GO:0005762 | mitochondrial large ribosomal subunit | Translation and Ribosome Biogenesis |
| GO:0003735 | structural constituent of ribosome | Translation and Ribosome Biogenesis |
| GO:0002181 | cytoplasmic translation | Translation and Ribosome Biogenesis |
| GO:0000981 | DNA-binding transcription factor activity, RNA polymerase II-specific | Translation and Ribosome Biogenesis |
| GO:0000463 | maturation of LSU-rRNA from tricistronic rRNA transcript (SSU-rRNA, 5.8S rRNA, LSU-rRNA) | Translation and Ribosome Biogenesis |
| GO:0000462 | maturation of SSU-rRNA from tricistronic rRNA transcript (SSU-rRNA, 5.8S rRNA, LSU-rRNA) | Translation and Ribosome Biogenesis |
| GO:0036297 | interstrand cross-link repair | DNA Replication Repair Recombination |
| GO:0006310 | DNA recombination | DNA Replication Repair Recombination |
| GO:0006284 | base-excision repair | DNA Replication Repair Recombination |
| GO:0006260 | DNA replication | DNA Replication Repair Recombination |
| GO:0006281 | DNA repair | DNA Replication Repair Recombination |
| GO:0003684 | damaged DNA binding | DNA Replication Repair Recombination |
| GO:0003697 | single-stranded DNA binding | DNA Replication Repair Recombination |
| GO:0003677 | DNA binding | DNA Replication Repair Recombination |
| GO:0000724 | double-strand break repair via homologous recombination | DNA Replication Repair Recombination |
| GO:0000723 | telomere maintenance | DNA Replication Repair Recombination |
| GO:0006457 | protein folding | Protein Folding |
| GO:0051539 | 4 iron, 4 sulfur cluster binding | Cofactor and Ion Binding |
| GO:0016226 | iron-sulfur cluster assembly | Cofactor and Ion Binding |
| GO:0008270 | zinc ion binding | Cofactor and Ion Binding |
| GO:0045271 | respiratory chain complex I | Mitochondrial and Cellular Respiration |
| GO:0045333 | cellular respiration | Mitochondrial and Cellular Respiration |
| GO:0042776 | proton motive force-driven mitochondrial ATP synthesis | Mitochondrial and Cellular Respiration |
| GO:0045259 | proton-transporting ATP synthase complex | Mitochondrial and Cellular Respiration |
| GO:0033617 | mitochondrial cytochrome c oxidase assembly | Mitochondrial and Cellular Respiration |
| GO:0032981 | mitochondrial respiratory chain complex I assembly | Mitochondrial and Cellular Respiration |
| GO:0015986 | proton motive force-driven ATP synthesis | Mitochondrial and Cellular Respiration |
| GO:0009060 | aerobic respiration | Mitochondrial and Cellular Respiration |
| GO:0008137 | NADH dehydrogenase (ubiquinone) activity | Mitochondrial and Cellular Respiration |
| GO:0006123 | mitochondrial electron transport, cytochrome c to oxygen | Mitochondrial and Cellular Respiration |
| GO:0006120 | mitochondrial electron transport, NADH to ubiquinone | Mitochondrial and Cellular Respiration |
| GO:0097542 | ciliary tip | Cellular Components / Localization |
| GO:0042645 | mitochondrial nucleoid | Cellular Components / Localization |
| GO:0005886 | plasma membrane | Cellular Components / Localization |
| GO:0005829 | cytosol | Cellular Components / Localization |
| GO:0005759 | mitochondrial matrix | Cellular Components / Localization |
| GO:0005737 | cytoplasm | Cellular Components / Localization |
| GO:0005743 | mitochondrial inner membrane | Cellular Components / Localization |
| GO:0005739 | mitochondrion | Cellular Components / Localization |
| GO:0005730 | nucleolus | Cellular Components / Localization |
| GO:0005654 | nucleoplasm | Cellular Components / Localization |
| GO:0005634 | nucleus | Cellular Components / Localization |
| GO:0001650 | fibrillar center | Cellular Components / Localization |

### 2.3. Biological interpretation using CURIE

To interpret the biological significance of the enriched functional categories, we employed CURIE (Version 2-3), an artificial intelligence tool designed to contextualize omics data through targeted literature mining and _116_ automated synthesis. CURIE was provided with the Gene Ontology (GO) terms found to be upregulated in bacterial pneumonia. Using these inputs, the platform systematically queried extensive scientific literature databases and generated narrative interpretations that integrated key molecular mechanisms, experimental findings, and current knowledge related to each biological process.

The Biofunctional app was used to interpret these ontologies and generate prompts that can be sent to AI models like Chat GPT or Gemini for further analysis. However, the app currently presents several limitations and errors and unfortunately the responses were not adequate. To address this, CURIE was developed.

CURIE isn’t directly integrated into the GANGO and BioFunctional platforms. Instead, once the GO terms of interest are selected, a list of these terms must be created. These GO terms can then be converted into a comma-separated list of description codes using an R script. This step is crucial for accurately interpreting the results and leveraging artificial intelligence for further analysis. A fully developed example of this methodology can be found in Rodriguez-Mena et al. (2025).

#### 2.3.1. The CURIE Framework: Foundations and Applications

To address the limitations of current tools, we are developing a custom AI solution in Python that leverages Retrieval-Augmented Generation (RAG) to enhance functional analysis capabilities. This effort is complemented by the creation of a confidential Python library, which integrates custom algorithms to improve the interpretation of biological datasets (Tarragó et al., 2025a; Rodriguez-Mena et al., 2025). Independent expert-based validation of CURIE-generated narratives has also been explored in other clinical domains, such as dermatology (Tardiolo et al., 2026). By combining RAG-based methods with advanced algorithmic frameworks, this approach aims to handle complex biological systems more effectively, laying a robust foundation for accurate and advanced data analysis.

Retrieval-Augmented Generation (RAG) is a major advancement in AI, leveraging deep learning techniques like transformer neural networks to process complex datasets (e.g., documents, bibliographies, and databases). RAG is particularly effective in answering complex queries by generating high-quality semantic representations.

RAG’s Two-Step Approach (Lewis et al, 2020):

Retriever: Identifies and ranks the most relevant documents using a probabilistic function p_n (z|x) with n parameters that returns (top-K truncated) distributions over text passages given a query x, selecting the top-kkk results for the query.

Generator: Produces coherent, context-aware responses by combining the retrieved documents with previously generated text, modeled as: p_θ (y_i│x,z,y_(1:i-1)) parametrized by θ that generates a current token based on a context of the previous i-1 tokens y_(1:i-1), the original input x and a retrieved passage z. p_θ (y_i│x,z,y_(1:i-) generates a comprehensive response (y) considering the query (x), retrieved document (z), and previously generated parts of the response (y1:i-1) that can be considered.

A key feature is RAG’s use of retrieved documents as a “knowledge grounding mechanism,” enabling accurate, context-driven responses. This makes RAG particularly valuable in fields like biological data interpretation, where precision and context are crucial.

Figure 4 illustrates the model’s architecture, which efficiently combines pre-trained retrievers and generators. The system uses MIPS (Maximum Inner Product Search) for efficient document ranking and a seq2seq generator to create coherent responses from sequential data. To effectively handle retrieval uncertainty, the model incorporates marginalization over documents. This entire process ensures high contextual relevance, resulting in generated responses that are both accurate and meaningful.

**Figure 4:**
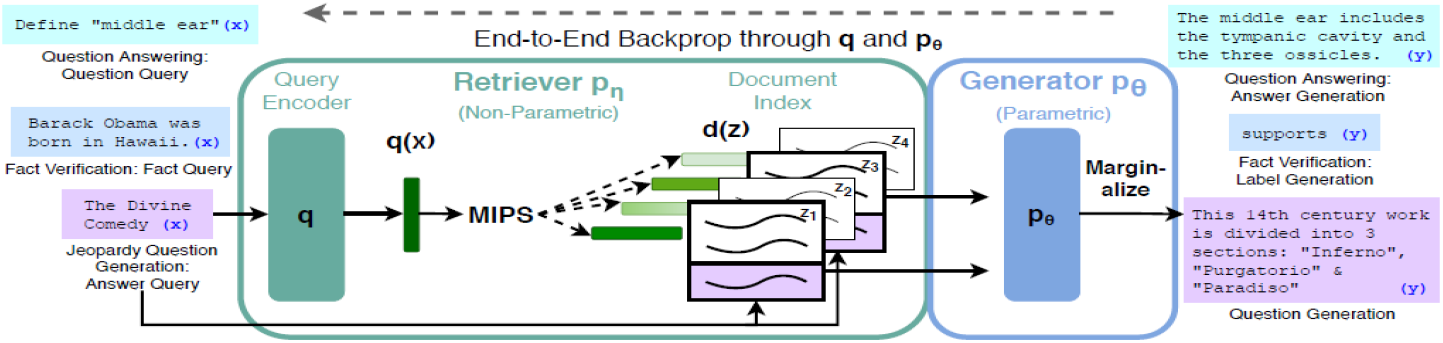
Example of RAG use. We combine a pre-trained retriever (Query Encoder + Document Index) with a pre-trained Sequence-to-sequence seq2seq model (Generator) and fine-tune end-to-end. For query x, we use Maximum Inner Product Search (MIPS) to find the top-K documents zi. For final prediction y, we treat z as a latent variable and marginalize over seq2seq predictions given different documents (from Lewis et al, 2020).

To train the retriever and generator end-to-end, the retrieved document is treated as a latent variable. Two models are proposed by Lewis et al. (2020) that marginalize over the latent documents in different ways to produce a distribution over generated text:

- RAG-Sequence: The model uses the same document to predict each target token.
- RAG-Token: Can predict each target token based on a different document.

RAG models combine a pre-trained retriever (DPR) with a pre-trained generator (BART). The retriever retrieves top-kkk relevant documents for a given query. The generator, treated as a latent variable, is then used to generate the response. RAG-Sequence uses the same document for the entire response, while RAG-Token can use different documents for each token. Both models are trained end-to-end, marginalizing over the retrieved documents.

### 2.3.2 Development of a Python Library to Train a RAG Model for Predicting Functional Analysis

A Python library has been developed to train a Retrieval-Augmented Generation (RAG) model aimed at predicting functional analysis interpretations from metagenomic data. This system was tested using data from the study, “A pilot study of the use of the oral and faecal microbiota for the diagnosis of ulcerative colitis and Crohn’s disease in a paediatric population” (Monleón et al., 2024). A detailed bioinformatic re-analysis of this same paediatric IBD cohort using the GANGO/BioFunctional pipeline, including KEGG and GO term interpretation, has since been reported in Tarragó et al. (2025b). The Python library leverages the RAG model to generate informed responses based on ontologies derived from metagenomic analyses. This AI has been installed on the research group’s servers and is currently undergoing validation (Rodriguez-Mena et al., 2025).

### 2.4. Availability of CURIE

CURIE (Version 5) is currently available at www.curieai.tech for non-profit purposes. A significant effort has been made to ensure that all aspects of the platform comply with European Union regulations, and it may not be used for commercial purposes. CURIE can be tested free of charge; however, intensive use entails computational and maintenance costs. We are actively working towards further developing this service to broaden its availability for external use, allowing the wider scientific community to leverage CURIE’s capabilities for enhanced functional analysis of biological datasets.

## 3. Results and discussion

### 3.1. DNA replication and repair

#### 3.1.1. DNA Repair Mechanisms

Individuals affected by bacterial pneumonia exhibit an upregulation of gene ontologies associated with DNA repair mechanisms when compared to healthy controls. Notably, an increase is observed in the general DNA repair term (GO:0006281), along with more specific up-regulations in the repair system, like in the interstrand cross-link repair (GO:0036297) pathway. Pathogen-associated molecular patterns (PAMPs), recognized by pattern recognition receptors (PRRs), initiate signaling cascades that modulate the host immune response, leading to the production of reactive oxygen species (ROS) as part of the body’s defense against the invading pathogens. ROS can cause DNA damage, which activates the base-excision repair pathway to fix the resulting lesions.

The interstrand cross-link repair is essential for resolving highly toxic DNA lesions in which both strands of the DNA double helix are covalently linked. Interstrand cross-links can be induced by endogenous metabolic byproducts, such as reactive oxygen species (ROS), or by exogenous agents including ionizing radiation (IR), ultraviolet (UV) radiation, and various chemical compounds (Hakem, 2008). These lesions obstruct DNA unwinding, thereby impairing fundamental cellular processes such as transcription and replication.

Also the base-excision repair (BER) (GO:0006284) mechanism is enhanced. It is responsible for correcting DNA damage caused by oxidation, deamination, and alkylation. The repair process is initiated by a DNA glycosylase that identifies and excises the damaged base, generating a basic site. This site is then processed through either short-patch or long-patch BER, which involve distinct sets of proteins to complete the repair (Krokan & Bjørås, 2013).

Additionally, double-strand break repair via homologous recombination (GO:0000724) is activated, which plays a crucial role in repairing DNA damage during mitosis and in facilitating chromosome pairing and genetic exchange during meiosis (Krejci et al., 2012).

These observations suggest that bacterial pneumonia induces a cellular environment characterized by elevated genotoxic stress, likely due to inflammation, oxidative damage, and pathogen-derived factors. In response, host cells selectively activate multiple DNA repair pathways to preserve genomic integrity.

#### 3.1.2. DNA Replication and Maintenance

In individuals affected by bacterial pneumonia, an upregulation is also observed in gene ontologies related to DNA replication and genome stability. DNA replication (GO:0006260) is enhanced, suggesting maybe an increased compensatory response to replenish damaged cells.

Telomere maintenance (GO:0000723) is also upregulated, possibly reflecting a cellular attempt to preserve chromosomal integrity under stress conditions, as telomeres are particularly sensitive to oxidative damage (Nguyen et al., 2025).

The activation of DNA recombination (GO:0006310) further supports a stress-adaptive response, as this process is essential for repairing damaged DNA, resolving replication fork stalling, and maintaining genome stability. DNA recombination refers to any process that results in the formation of a new genotype through the reassortment of genetic material, producing gene combinations that differ from those found in the parent organisms Together, the upregulation of these pathways indicates a broader cellular effort to sustain genomic function and viability during bacterial infection.

### 3.2. DNA Binding Functions

The general upregulation in DNA binding functions further reinforces the earlier findings. DNA binding (GO:0003677) is a broad category encompassing proteins that interact with DNA to regulate replication, repair, transcription, and structural maintenance. The upregulation of damaged DNA binding (GO:0003684) indicates that cells are actively recognizing and responding to lesions in the DNA, likely caused by inflammation and oxidative stress. An example of this is the UV-damaged DNA-binding protein (UV-DDB), which binds to DNA lesions caused by ultraviolet (UV) radiation (Sugasawa, 2009). Additionally, the increase in ssDNA binding (GO:0003697) reflects a heightened need to stabilize and protect exposed ssDNA intermediates, which arise during replication, repair, and recombination processes. Together, these changes support the idea of an intensified DNA damage response and enhanced engagement of repair and replication machinery in the infected state.

### 3.3. Transcription and RNA Processing

#### 3.3.1. Transcription and Transcriptional Regulation

In bacterial pneumonia, the upregulation of gene ontologies related to transcriptional activity suggests a global enhancement of RNA synthesis and gene expression regulation. The enhancement in DNA-directed RNA polymerase activity (GO:0003899) reflects the up-regulation in the core enzymatic function responsible for transcribing DNA into RNA, a fundamental process required for cellular response and adaptation (Hsieh & Borger, 2023). The upregulation of the RNA polymerase III complex (GO:0005666) indicates increased transcription of small RNAs, such as tRNAs, 5S rRNA and U6 snRNA (Li et al., 2021). The enrichment of RNA polymerase II cis-regulatory region sequence-specific DNA binding (GO:0000978) refers to the interaction of transcription factors with specific upstream regulatory DNA sequences—known as recognition sequences or binding sites—located on the same DNA molecule as the gene being transcribed. These interactions regulate the activity of RNA polymerase II, the enzyme responsible for synthesizing messenger RNA in eukaryotic cells. This is consistent with the observed upregulation in regulation of transcription by RNA polymerase II (GO:0006357), highlighting a coordinated control of gene expression programs in response to infection-induced stress.

#### 3.3.2. RNA Processing and Maturation

The upregulation of gene ontologies related to RNA processing and maturation indicates an enhanced co-transcriptional and post-transcriptional regulatory activities, coherent with the up-regulation in transcription. In particular, the activation of RNA splicing (GO:0008380) and specifically mRNA splicing via the spliceosome (GO:0000398) suggests increased processing of pre-mRNAs into mature transcripts, a critical step for proper gene expression. Pre-mRNA processing, including 5′-end capping, splicing, editing, and polyadenylation, is a series of coordinated and largely cotranscriptional events that ensure the high fidelity and functional diversity characteristic of eukaryotic gene expression. Mechanisms such as alternative splicing and RNA editing allow a relatively limited number of genes to produce a wide range of protein isoforms, while alternative 3′-end formation contributes to transcript-specific regulation of localization, translation efficiency, and stability (Poulos et al., 2011).

The enhancement in pre-mRNA processing and splicing is further supported by the up-regulation shown by the GO associated with spliceosomal components and assembly. The spliceosome, a ribonucleoprotein (RNP) complex composed of five snRNPs and numerous associated proteins, catalyzes pre-mRNA splicing. During its assembly and catalytic cycle, the spliceosome forms and dynamically rearranges complex RNA-RNA and RNP interactions to precisely align the reactive groups of the pre-mRNA for splicing. Most eukaryotes possess two distinct spliceosomes: the U2-dependent spliceosome, which mediates the removal of common U2-type introns, and the U12-dependent spliceosome, a less abundant complex found in a subset of eukaryotes that processes the rare U12-type class of introns. The U2-dependent spliceosome is composed of the U1, U2, U5, and U4/U6 snRNPs, along with numerous non-snRNP proteins that assist in its assembly and function. In contrast, the U12-dependent spliceosome includes U11, U12, U5, and U4atac/U6atac snRNPs as its core components (Will & Lührmann, 2011).

While the U2-type complex is responsible for splicing the vast majority of canonical introns, the U12-type spliceosome targets a rare class of non-canonical introns (U12-type), which are often found in genes with essential regulatory roles.

In the case of individuals affected by bacterial infection, the general enhancement in spliceosomal complex (GO:0005681) is accompanied by the upregulation of the U2-type spliceosomal complex (GO:0005684) and U2-type pre catalytic spliceosome (GO:0071005). This reflects heightened activity in the assembly of functional splicing intermediates, while the enrichment of the catalytic step 2 spliceosome (GO:0071013) suggests that the full splicing cycle is actively completed. Interestingly, the upregulation of the U12-type spliceosomal complex (GO:0005689) indicates that even the minor spliceosome is engaged during infection.

The upregulation of GO terms associated with spliceosomal components indicates an overall increase in spliceosome assembly, activation, and catalytic activity. The enrichment of spliceosomal snRNP assembly (GO:0000387) suggests enhanced biogenesis and recruitment of small nuclear ribonucleoproteins (snRNPs), a critical early step for the formation of functional spliceosomes. Specific components of the U2-dependent spliceosome, including U1 snRNP (GO:0005685), U2 snRNP (GO:0005686), and U5 snRNP (GO:0005682), are all upregulated, pointing to a robust activation of the major splicing machinery. The presence of increased levels of the U4/U6 × U5 tri-snRNP complex (GO:0046540) further confirms efficient spliceosome assembly, as this complex is essential for the transition from early to catalytically active splicing stages.

Additionally, we can observe the upregulation of tRNA processing (GO:0008033) and rRNA processing. In eukaryotic cells, distinct DNA-dependent RNA polymerases—Pol I, Pol II, and Pol III—are responsible for transcribing different classes of RNA. Specifically, Pol I synthesizes precursor ribosomal RNA (pre-rRNA), Pol II transcribes precursor messenger RNA (pre-mRNA), and Pol III produces precursor transfer RNA (pre-tRNA). The processing pathways for these pre-RNAs differ substantially in both timing and subcellular localization. For instance, pre-mRNA undergoes co-transcriptional processing, including capping, splicing, polyadenylation, and nuclear export, all of which are tightly coupled to transcription. In contrast, tRNA biogenesis is largely post-transcriptional, occurring across multiple specialized subcellular compartments (Hopper & Nostramo, 2019).

Also, the rise in tRNA binding (GO:0000049) and tRNA methylation (GO:0030488), a modification essential for tRNA stability and accurate decoding, further supports a tightly regulated post-transcriptional response aimed at maintaining translational fidelity under conditions of infection-induced stress.

### 3.4. Translation and Ribosome Biogenesis

#### 3.4.1 Translation Process

Protein synthesis is a fundamental process that has been remarkably conserved across evolution. It proceeds from mRNA templates and results in the production of polypeptide chains. The core mechanism of translation is consistent across all domains of life: it occurs on ribosomes, with transfer RNAs (tRNAs) acting as adaptors that match specific codons on the mRNA to their corresponding amino acids. Protein synthesis relies on the coordinated function of three classes of RNA molecules: messenger RNA (mRNA), transfer RNA (tRNA), and ribosomal RNA (rRNA), along with a range of protein factors that facilitate the various stages of translation (Cooper, 2000).

The upregulation of GO involved in the translation process suggests a marked enhancement of protein synthesis in individuals affected by bacterial pneumonia. Translation (GO:0006412) and cytoplasmic translation (GO:0002181) are both significantly enriched, indicating a global activation of cytosolic protein production. This is accompanied by the upregulation of mitochondrial translation (GO:0032543), reflecting increased mitochondrial activity, likely to meet elevated energy demands during infection.

In parallel, the enrichment of translational initiation (GO:0006413) and the eukaryotic 48S preinitiation complex (GO:0033290) highlights an active initiation phase of translation. Furthermore, the upregulation of cytosolic ribosome GO (GO:0022626) points to an increased availability of ribosomal machinery necessary for sustained translation.

During translation, each of the 20 amino acids must be matched with its corresponding codon on the mRNA, a task carried out by tRNAs, which act as adaptors. Accurate protein synthesis depends on both proper codon-anticodon base pairing and the correct attachment of each amino acid to its corresponding tRNA. This attachment is catalyzed by a family of enzymes known as aminoacyl-tRNA synthetases (Cooper, 2000).

The elevation of tRNA aminoacylation for protein translation (GO:0006418) and tRNA binding, whose GO was already discussed in the previous section, further suggests a boost in translational accuracy and efficiency, as these processes are critical for charging tRNAs with their respective amino acids and ensuring correct codon-anticodon pairing during elongation. Altogether, these changes reflect a coordinated cellular response aimed at increasing the capacity for protein synthesis under conditions of infection-induced stress. In fact, the observed enhancement in translation may reflect the host cell’s need to rapidly produce proteins essential for mounting and sustaining an effective response to bacterial infection. During conditions of stress and inflammation, cells must upregulate the synthesis of a wide array of proteins, including cytokines, chemokines, and components of the immune signaling network. This increased demand requires not only higher activity of the core translational machinery, such as ribosomes and translation initiation complexes, but also enhanced processing and charging of tRNAs to maintain accuracy and efficiency in protein production (Argüello et al., 2015).

#### 3.4.2. Ribosome Assembly and Biogenesis

Protein synthesis in eukaryotes is mediated by the ribosome, a large ribonucleoprotein complex composed of a small and a large subunit. During translation, the small subunit decodes the mRNA sequence while the large subunit catalyzes peptide bond formation, coupling mRNA decoding with polypeptide chain elongation (Klinge & Woolford, 2019).

The upregulation of gene ontologies related to ribosomal subunits and biogenesis further indicates a significant boost in the host’s translational capacity during bacterial pneumonia.

First of all, we observe the up-regulation of the general term Ribosome (GO:0005840), together with its structural constituent (GO:0003735). Specifically, both the cytosolic large (GO:0022625) and small (GO:0022627) ribosomal subunits are enhanced, suggesting an increased availability of functional ribosomes for protein synthesis.

Ribosome biogenesis in eukaryotic cells is a highly complex and tightly regulated process that involves the coordinated activity of all three nuclear RNA polymerases. It begins in the nucleolus, where RNA polymerase I transcribes the 47S pre-rRNA, which is processed into the 18S, 5.8S, and 28S rRNAs. In parallel, RNA polymerase III independently transcribes 5S rRNA, while RNA polymerase II transcribes ribosomal protein genes, mRNAs are translated in the cytoplasm. The resulting ribosomal proteins are imported into the nucleus and assembled with rRNAs to form the 90S processome, which is then split into pre-40S and pre-60S subunits. These subunits undergo further maturation in the nucleoplasm and cytoplasm before becoming fully functional ribosomes (Ni & Buszczak, 2023).

The upregulation of ribosomal small subunit biogenesis (GO:0042274) and ribosomal large subunit biogenesis (GO:0042273), along with their respective precursors, small-subunit processome (GO:0032040) and pre ribosome, large subunit precursor (GO:0030687), reflects an active ribosome assembly process, essential for sustaining high rates of translation under stress conditions.

Additionally, the increased expression of mitochondrial ribosome (GO:0005761) and its large (GO:0005762) and small (GO:0005763) subunits highlights the parallel upregulation of mitochondrial translation. Mitochondria are semi-autonomous organelles that possess their own genetic system and are capable of independently replicating, transcribing, and translating their mitochondrial DNA. An up-regulation in the GO associated with their translation, likely supports elevated energy production and mitochondrial protein synthesis during the host immune response.

Collectively, these changes suggest a tightly regulated expansion of both cytosolic and mitochondrial ribosomal machinery to meet the cellular demands imposed by infection.

Associated with the Ribosomal assembly and biogenesis, we observe an upregulation of gene ontologies associated with rRNA maturation and binding. This highlights an increase in the production and processing of ribosomal RNA, which is essential for assembling functional ribosomes. Specifically, the enrichment of maturation of SSU-rRNA from tricistronic rRNA transcript (GO:0000462) and maturation of LSU-rRNA from tricistronic rRNA transcript (GO:0000463) indicates active processing of the 47S pre-rRNA transcript into the small (18S) and large (5.8S and 28S) ribosomal subunit rRNAs, a critical step in ribosome biogenesis. Additionally, the upregulation of maturation of SSU-rRNA (GO:0030490) reflects focused efforts on assembling the small ribosomal subunit. The increased expression of rRNA binding (GO:0019843) further supports this enhanced biogenetic activity, as numerous ribosome assembly factors and ribosomal proteins interact directly with rRNA to facilitate its correct folding, processing, and incorporation into ribosomal subunits.

### 3.5. Protein folding

For polypeptides to become functional, they must fold into specific three-dimensional structures, and in many cases, multiple polypeptide chains must assemble into higher-order complexes to form active proteins (Cooper, 2000). The observed upregulation of protein folding (GO:0006457) completes the pathway of protein synthesis by ensuring that newly translated polypeptides acquire their correct three-dimensional structure. The increased expression of folding-related genes suggests that, in response to bacterial infection, host cells not only accelerate translation but also reinforce the systems required to process and maintain protein quality. This response likely involves the activation of molecular chaperones and folding enzymes that assist in the proper folding of nascent chains, prevent aggregation, and help refold stress-damaged proteins. Together with the upregulation of translation and ribosome biogenesis pathways, the enhancement of protein folding machinery reflects a comprehensive translational and proteostatic response aimed at maintaining cellular function and resilience under infectious stress.

### 3.6. Mitochondrial Energy Metabolism and Respiratory Chain Assembly

#### 3.6.1. Electron Transport Chain (ETC) Function

The electron transport chain (ETC) is a series of four protein complexes located in the inner mitochondrial membrane that couple redox reactions to the generation of an electrochemical gradient. This gradient drives the production of ATP through oxidative phosphorylation. In the context of cellular respiration, electrons are derivedfrom the breakdown of organic molecules such as glucose, and their transfer through the ETC releases energy that is harnessed to power ATP synthesis.

Aerobic cellular respiration consists of three main stages: glycolysis, the citric acid (Krebs) cycle, and oxidative phosphorylation. In the final stage, oxidative phosphorylation, the NADH and FADH^2^ generated from earlier steps, donate electrons to the electron transport chain (ETC). As electrons flow through the ETC via redox reactions, energy is released and used to pump protons across the membrane, generating a proton gradient (Ahmad, Wolberg, & Kahwaji, 2023).

The upregulation during bacterial pneumonia of gene ontologies associated with the electron transport chain (ETC) indicates a significant enhancement of mitochondrial oxidative metabolism in response to bacterial infection. Key processes such as electron transport from NADH to ubiquinone (GO:0006120) and from cytochrome c to oxygen (GO:0006123) suggest increased activity of complex I and complex IV, two critical components of the ETC that drive the transfer of electrons along the mitochondrial inner membrane. The enrichment of NADH dehydrogenase (ubiquinone) activity (GO:0008137) and respiratory chain complex I (GO:0045271) further confirms this elevation in mitochondrial respiration. These processes contribute directly to aerobic respiration (GO:0009060) and overall cellular respiration (GO:0045333), which are essential for generating the energy required to support elevated biosynthetic activity, stress responses, and immune defense mechanisms.

#### 3.6.2. ATP Synthesis and Proton Motive Force

The majority of ATP synthesis takes place during cellular respiration in the mitochondrial matrix, yielding approximately 32 ATP molecules per oxidized glucose molecule. This ATP serves as the primary energy currency of the cell and is consumed in various essential processes, including ion transport, muscle contraction, nerve impulse propagation, substrate-level phosphorylation, and biosynthetic reactions (Dunn & Grider, 2023).

The upregulation of gene ontologies related to ATP synthesis and the proton motive force, the energy generated by the transfer of protons or electrons across an energy-transducing membrane and that is typically used to drive processes such as ATP synthesis, reflects increased mitochondrial activity aimed at meeting the elevated energy demands induced by bacterial infection. Proton motive force-driven mitochondrial ATP synthesis (GO:0042776) and proton motive force-driven ATP synthesis (GO:0015986) indicate enhanced coupling of oxidative phosphorylation to ATP production.

These processes rely on the proton-transporting ATP synthase complex (GO:0045259), which utilizes the electrochemical gradient generated by the electron transport chain to catalyze the phosphorylation of ADP to ATP. The enrichment of these pathways suggests that host cells are maximizing energy output through oxidative metabolism, likely to sustain increased protein synthesis, stress responses, and other biosynthetic demands during infection.

#### 3.6.3. Respiratory Complex Assembly

The upregulation of gene ontologies involved in respiratory complex assembly suggests an active remodeling or reinforcement of mitochondrial oxidative capacity in response to bacterial infection. Specifically, mitochondrial respiratory chain complex I assembly (GO:0032981) and mitochondrial cytochrome c oxidase assembly (GO:0033617) refer to the formation of complex I and complex IV, respectively. Both the activities associated with these complexes, electron transport from NADH to ubiquinone and electron transport from cytochrome c to oxygen, are seen to be up-regulated. Proper assembly of these multi-subunit complexes is critical for maintaining efficient electron flow, minimizing reactive oxygen species production, and sustaining ATP generation through oxidative phosphorylation.

#### 3.7. Cofactor and ion binding

The upregulation of gene ontologies related to cofactor and ion binding suggests an increased demand for metalloenzyme function and mitochondrial redox activity in response to bacterial infection.

First of all, an upregulation in individuals affected by bacterial pneumonia in zinc ion binding (GO:0008270) is observed. Zinc (Zn) is a versatile trace element essential for virtually all aspects of eukaryotic cell biology. Zinc ions play a crucial role in various cellular processes in eukaryotic organisms, including signal transduction, gene expression, DNA repair and apoptosis regulation (Costa, Sarmento-Ribeiro, & Gonçalves, 2023). They are also involved in numerous metabolic pathways and are essential for the proper functioning of the immune system.

The enrichment of iron-sulfur cluster assembly (GO:0016226) and 4 iron, 4 sulfur cluster binding (GO:0051539) reflects heightened mitochondrial activity, as these clusters serve as key cofactors in redox reactions and are critical for the function of electron transport chain complexes, including complex I and II (Rouault & Tong, 2008). This thus supports the up-regulations in the GO associated with the ETC pathway. In fact, their upregulation likely supports enhanced respiratory activity, oxidative phosphorylation, but also DNA repair and genome integrity. All these processes are vital for maintaining cellular homeostasis under the metabolic and oxidative stress conditions triggered by infection.

### 3.8. Cellular Localization and Subcellular Compartmentalization

#### 3.8.1. Nuclear Compartments

The upregulation of GO terms associated with nuclear compartments suggests an intensification of nuclear activity in response to bacterial infection. The nucleus (GO:0005634) and nucleoplasm (GO:0005654), which are in fact up-regulated, are central to transcriptional regulation, DNA replication, and repair. In parallel, the enrichment of the nucleolus (GO:0005730)—a subnuclear structure involved in rRNA transcription and ribosome biogenesis— indicates increased protein synthesis demand. Similarly, the fibrillar center (GO:0001650), which is part of the nucleolar architecture where rDNA transcription is initiated, further supports heightened ribosomal RNA production and nucleolar function under stress conditions.

#### 3.8.2. Mitochondrial Compartments

The upregulation of mitochondrial-related GO terms reflects enhanced mitochondrial activity and metabolic adaptation during infection. The mitochondrion (GO:0005739), as a whole, shows increased involvement, accompanied by specific enrichment in the mitochondrial inner membrane (GO:0005743) and matrix (GO:0005759), key sites of oxidative phosphorylation and metabolic enzyme activity. The mitochondrial inner membrane houses the protein complexes of the electron transport chain (ETC) and ATP synthase. Its upregulation further supports the intensification of the oxidative phosphorylation pathway, leading to greater ATP production. At the same time also, the matrix indicates an increase in the TCA cycle. In fact, it contains enzymes of this and other metabolic pathways, as well as mitochondrial DNA and ribosomes.

The up-regulation of mitochondrial nucleoid (GO:0042645), which contains mtDNA and its associated transcription/replication machinery, indicates a general increase in these functions, likely to supply the components needed for expanded mitochondrial capacity and to support the increased biosynthesis of ETC proteins encoded by the mitochondrial genome.

#### 3.8.3. Cytoplasmic and Peripheral Structures

The increased expression of GO terms related to cytoplasm (GO:0005737) and cytosol (GO:0005829) points to an overall elevation in cytoplasmic processes such as translation, signaling, and protein transport. As the primary site for a wide array of biochemical reactions, including protein synthesis, signaling cascades, and vesicle trafficking, increased activity in the cytoplasm suggests broad upregulation of essential cellular functions. This is supported also by the up-regulation of its soluble part, the cytosol. In fact, it is a key hub for metabolic pathways, mRNA translation, and signal transduction.

The upregulation of the plasma membrane (GO:0005886) implies increased receptor expression and activity, particularly for pattern recognition receptors (PRRs), cytokine receptors, and antigen-presenting complexes. These components are central to detecting pathogens, initiating downstream signaling, and orchestrating communication between immune cells. Additionally, the enrichment at the ciliary tip (GO:0097542), a specialized structure involved in signal reception and trafficking of signaling molecules in cells with primary cilia.

### 3.9. Limitations

This study should be interpreted as a proof-of-concept illustrating the integration of AI-based literature mining into functional transcriptomic analysis, rather than a validated biological finding. Several limitations should be noted. First, the narrative interpretations generated by CURIE were not subjected to quantitative validation against an independent gold standard or expert-blinded review; their biological plausibility was assessed qualitatively by the authors. Second, several of the mechanistic links proposed between enriched GO terms and hostpathogen interaction (e.g., links to oxidative stress, PAMP recognition) are drawn from the general literature retrieved by CURIE and should be considered as plausible hypotheses rather than directly demonstrated findings in this dataset. Third, CURIE is currently under active development and validation by the BIOST3 research group, and its outputs may change as the underlying retrieval and generation components are refined. Future work should include systematic benchmarking of CURIE-generated interpretations against expert annotation and, where possible, experimental validation of the proposed mechanisms. A first step in this direction has been taken in a pilot expert-audit study assessing dermatologist concordance with CURIE-AI-generated Gene Ontology narratives in psoriasis (Tardiolo et al., 2026), which found moderate-to-good agreement (Fleiss’ κ = 0.466; ICC up to 0.812 for single-GO reports) but reduced reliability for grouped ontology clusters, underscoring the need for the same kind of expert-centred validation illustrated qualitatively in the present study.

## 4. Conclusions

In conclusion, our study successfully leveraged CURIE, an advanced AI platform designed for mining biological literature and generating contextual annotations, to significantly enhance the interpretation of complex transcriptomic data. By systematically cross-referencing Gene Ontology (GO) terms with extensive scientific databases, CURIE moves beyond traditional enrichment outputs, providing narrative explanations that integrate key molecular mechanisms, experimental evidence, and relevant publications. This capability proved invaluable in contextualizing the systemic molecular responses observed in bacterial pneumonia, offering a deeper, literature-supported understanding of the involved biological processes.

CURIE’s utility is evident in its ability to provide a more holistic understanding of host-pathogen interactions at the transcriptomic level by combining classical statistical enrichment with AI-based interpretation. The AI platform has been successfully installed on our research group’s servers and is currently undergoing validation, including formal expert-concordance studies in other clinical domains (Tardiolo et al., 2026) and additional applications to microbiome-based functional analysis in paediatric inflammatory bowel disease (Tarragó et al., 2025b). Integration of its methodology into the GANGO and BioFuncional platforms is planned for future development, with the aim of providing more precise and automated interpretations of metagenomic data. Ultimately, the integration of CURIE’s AI-supported insights with expert biological judgment facilitated a thorough and well-reasoned analysis, demonstrating the powerful potential of this novel approach in advancing functional analysis.

Finally, the spatial organization of this response is evident in the upregulation of GO terms defining nuclear, mitochondrial, cytoplasmic, and peripheral compartments, indicating a coordinated compartmentalization of the infection-induced cellular activity.

Altogether, these findings indicate that bacterial pneumonia provokes a systemic and coupled cell response dominated by enhanced metabolic, transcriptional, and translational activity. Such alterations probably function to enhance defense against infection, repair bacterial pathogenic injury, and preserve cell structure despite infectious stress.

## Author Contributions

The authors acknowledge their contributions to this paper as follows: Study conception and design: AR, AMG; Data collection: AR, XT; Analysis and interpretation of results: XT, GC, JM, VM, YT; Manuscript draft preparation: AMG, AR, GC. All authors reviewed the results and approved the final version of the manuscript.

### Funding

This research received no external funding. CURIE is registered by the University of Barcelona (2026).

## Acknowledgments

During the preparation of this manuscript, the artificial intelligence tool CURIE (version 5), developed by the BIOST3 research group (University of Barcelona), was used. Access to the tool was through a specific controlled environment provided by the BIOST3 group in September 2026. Its use was limited exclusively to linguistic, editorial, and bibliographic support tasks: composition and improvement of writing, spelling and grammar checks, and assistance in the organization and formatting of references. The tool did not perform any analytical work and did not participate in the study design, the collection, processing, or interpretation of the experimental data, which are the sole result of the author’s work and that of the collaborators cited in the acknowledgments. All content generated or suggested by the AI has been reviewed, corrected, and validated by the author, who assumes full responsibility for the accuracy, integrity, and originality of the final text.

## Declaration on the Use of Artificial Intelligence

This study made use of the artificial intelligence tool CURIE (version 5 and previous), developed by the BIOST3 research group (University of Barcelona), in two distinct and clearly delimited contexts. First, as described in Section 2.3 (‘Biological interpretation using CURIE’) and discussed throughout Section 3 (‘Results and discussion’), CURIE was used as a research instrument of this study: it was provided with the list of significantly enriched Gene Ontology (GO) terms and employed to retrieve and synthesize relevant scientific literature, generating narrative explanations that linked molecular mechanisms, experimental evidence, and publications to each biological process. This use constitutes a core part of the methodology under evaluation in this manuscript and is reported, with its limitations, as part of the study’s findings; it is not a tool used merely to assist in the writing of the manuscript. Second, independently of the above, the same tool was used during the preparation of the manuscript text, limited exclusively to linguistic, editorial, and bibliographic support tasks: improvement of writing style, spelling and grammar checks, and assistance in the organization and formatting of references. The tool did not perform any analytical work and did not participate in the study design, data collection, data processing, or in generating the scientific conclusions of this study, which remain the sole responsibility of the authors. All content generated or suggested by CURIE, in either of the two uses described above, has been reviewed, corrected, and validated by the authors, who assume full responsibility for the accuracy, integrity, and originality of the final text and for the scientific validity of the interpretations presented.

## Data Availability Statement

The RNA-Seq dataset analyzed in this study is publicly available in the Gene Expression Omnibus (GEO) repository under accession number GSE161731. The list of GO terms used as input for CURIE, along with the GANGO/BioFunctional output tables underlying Table 1, are available from the corresponding author upon reasonable request.

